# Forecasting Preventive Dental Quality Measures

**DOI:** 10.1101/2021.08.29.21262443

**Authors:** Radhakrishnan Nagarajan, Aloksagar Panny, Megan Ryan, Scott Murphy, Marko Vujicic, Gregory Nycz

**Author notes:** **Correspondence to:** Radhakrishnan Nagarajan, Center for Oral-Systemic Health, Marshfield Clinic Research Institute, Marshfield Clinic Health System, Marshfield, WI.

## Abstract

Dental quality measures objectively measure the efficiency and performance of dental providers and organizations. While these measures in conjunction with established benchmarks are used routinely for self-assessment, forecasting them ahead of time in a data-driven and evidence-based manner has the potential to assist in assessing future dental treatment needs, oral disease burden, care utilization patterns, and strategic decision making for sustained performance improvement complementing traditional descriptive visualization dashboards. The present study modeled the temporal trends of four key preventive dental quality measures related to caries prevention (Adult New Caries, Sealants (6-9yrs.), Sealants (12-15yrs.), Fluoride Varnish) sampled monthly from (Dec. 2010 to July 2017) averaged across ten Family Health Center Dental Centers (FQHC), Wisconsin, using auto-regressive integrated moving average time series models. Five-month ahead forecasts along with their 95% confidence levels and mean absolute percentage error were determined across the four measures (Adult New Caries: 1.8%, Sealants (6-9yrs.): 0.90%, Sealants (12-15yrs.): 0.30%, Fluoride Varnish: 0.15%). Model diagnostics revealed auto-regressive integrated moving average models to sufficiently capture the temporal patterns of these measures and the forecast estimates of Adult New Caries and Sealants (12-15yrs.) revealed the need for increased efforts for improved preventive care utilization. Forecasting preventive dental quality measures can provide insights into expected treatment needs ahead of time and can assist in optimal resource and staff allocation with potential to prescribe suitable interventions to shift the trajectory from predicted outcomes to desired outcomes in a targeted manner. While the present study investigated organization level preventive dental quality measures, the time series approach presented is as such generic and expected to translate across similar settings.

## Introduction

The national health care expenditure of United States grew by 4.6% in 2019 accounting for 17.7% of the Gross Domestic Product^1^. It is projected to grow at an average annual rate of 5.4% from 2019-2028 reaching a total of $6.2 trillion^1^. Increasing cost of care demand improved care quality for optimal performance of healthcare organizations. The Triple Aim defined by the Institute for Healthcare Improvement emphasizes the need to improve population health, experience of care while minimizing per capita cost of care^2^. Current emphasis on transitioning from fee for service to value-based models and overwhelming evidence on the bi-directional nexus between oral and systemic health^3,4^, especially demand measuring the performance objectively in order to achieve the triple aim^2,3,5^. The Physician Group Practice **(PGP)** demonstration project by the Center for Medicare and Medicaid Services (**CMS**) was the first legislatively mandated pilot pay-for-performance initiative that incentivized quality of care^3,4^. The performance of the participating entities was assessed using a set of quality measures drawn from CMSs Doctor’s Office Quality project. Marshfield Clinic Health System was one of the ten centers invited to participate in the PGP demonstration. Following the IOM reports on the quality improvement^5,6^, several national initiatives were also proposed to improve quality of care. This includes the Affordable Care Act and creation of Accountable Care Organizations to improve the quality of care for Medicare fee-for-service beneficiaries^7^. Financial incentives under these initiatives have especially emphasized the need to lower costs while meeting quality^8^.

Lack of quality measures in dentistry had been identified as a barrier by the Institute of Medicine in improving oral health outcomes and reducing oral health disparities^9–13^. The American Dental Association upon request by CMS established the Dental Quality Alliance (**DQA**) in 2008 for developing standardized dental quality measures (**DQM**’s) in an effort to improve oral health, patient care and safety through a consensus building approach^14,15^. The Measures Development and Maintenance Committee of DQA developed DQMs that are meaningful, transparent and collaborative^14^. A recent systematic review summarizes the evolution of DQMs^16^. The Family Health Center of Marshfield comprising of 10 dental centers (**FHC**), were early adopters of DQMs as a part of their dental quality improvement initiative. They implemented key clinical and operational DQMs in a web-based tool format (Dental Quality Analytics Dashboard)^17^ adopted by dental providers, staff and administrators for quality improvement at FHC as well as achieving value-based care in a predominantly Medicaid population in rural Wisconsin^18^. A steering committee including clinical and operational leadership, Dental and Medical Directors, and stakeholders were established to guide the successful implementation and adoption of the dashboard. The dashboard was used to report and monitor practice-level changes. The dashboard also facilitated comparison of clinical and operational DQM’s defined by the National Network for Oral Health Access (NNOHA) and Health Resources and Services Administration (HRSA) at both provider and organizational level. A recent study^17^ provides a detailed description of the architecture and measures implemented in the dashboard at FHC. More recently, FHC was also one of the first dental organization to publicly report (2020) dental quality measures (caries risk assessment in children, ongoing care in adults with periodontitis, topical fluoride application in high-risk children) along with four regional members through the Wisconsin Collaborative for Healthcare Quality as well as develop oral-systemic quality measures that use integrated medical-dental electronic health records. Adoption of quality measures at FHC has also resulted in marked improvement in preventive care utilization and value-based dental care delivery in a predominantly Medicaid population^18^.

While the past implementation^17^ had focused primarily on the visualization of DQM temporal trends using dashboards (i.e. descriptive analytics), the present study marks the shift from descriptive to predictive analytics that forecasts the measures using time series models. Such forecasts in turn are expected to provide preliminary cues for targeted interventions so as to improve performance across select measures and shift the trajectory from predicted outcomes to desired outcomes in a timely manner. More specifically, the present study investigates forecasting four preventive DQMs with a focus on caries prevention (Adult New Caries, Sealants (6-9yrs), Sealants (12-15yrs), Fluoride Varnish) averaged across the ten FHC dental centers from (Dec. 2010 to July 2017) using time series models. Dental caries has been associated with significant economic burden ^16,19,20^. National Center for Statistics 2018 report indicates 16.9% of children (5-19 yrs.) and 31.6% of adults (20-44 yrs.) had untreated dental caries in 2013-2016. CDCs 2017 Morbidity Mortality Weekly Report (MMWR)^21^ also emphasizes the prevalence of untreated dental caries in children and adolescents. More importantly, the report emphasized an increase in dental caries with age (6.1% in 6-11yrs. and 14.5% in 12-15 yrs. 2011-2014). Evidence based clinical guidelines for reducing the risk of dental caries among high-risk individuals recommend placement of sealants on the pits and fissures of occlusal surfaces in both primary and permanent molars in children and adolescents^22^. Professional application of topical fluoride varnish is also recommended for preventing dental caries^23^. DQMs have the ability to objectively assess caries prevention in children^24–27^. While preventive dental quality measures are routinely used in conjunction with established benchmarks, forecast models of these measures from electronic health records of a given population has the potential to reveal patterns in a data-driven and evidence-based manner. These in turn can assist in strategic decision making and sustained performance improvement with a direct impact on outcomes^5^. More importantly, such forecasts can assist in assessing preventive dental treatment needs and oral disease burden ahead of time minimizing aggressive disease. Forecast models are also expected to assist in targeted and optimal resource and staff allocation to areas of immediate need, assess preventive care utilization in a given population and develop polices and interventions ahead of time at the system-level that impacts various stakeholders (i.e. patients, providers, policymakers)^28^ for improved outcome.

## Materials and Methods

### Data Description

The four preventive DQMs investigated in the present study were obtained from the dental electronic health record and stored in an Enterprise Data Warehouse at the Marshfield Clinic Health System and were in the form of a time series, **Table 1**. The study was approved by the Marshfield Clinic Research Institute’s Institutional Review Board. It is important to note that only aggregate data at the organization level is investigated in the present study. The time period (Dec. 2010 to July 2017) was chosen since the monthly data of the four DQMs were available across all the ten FHC dental centers during this time frame. Their values were subsequently averaged to generate the averaged DQM time series data of the FHC investigated in the present study. Since the time series represent monthly samples, there were totally (N = 80) time points corresponding to 80 months. The first (N = 75) time points were used to model the DQM time series and its predictive ability and performance was subsequently demonstrated on the remaining (N = 5) time points (i.e. five month ahead forecast) corresponding to months March 2017 March to July 2017. Brief description of the four preventive DQMs is enclosed below. The numerator and denominator used in computing these DQMs along with the benchmarks are also enclosed in **Table 1**. A more detailed description can be found in our recent study^17^.

**Table 1.**
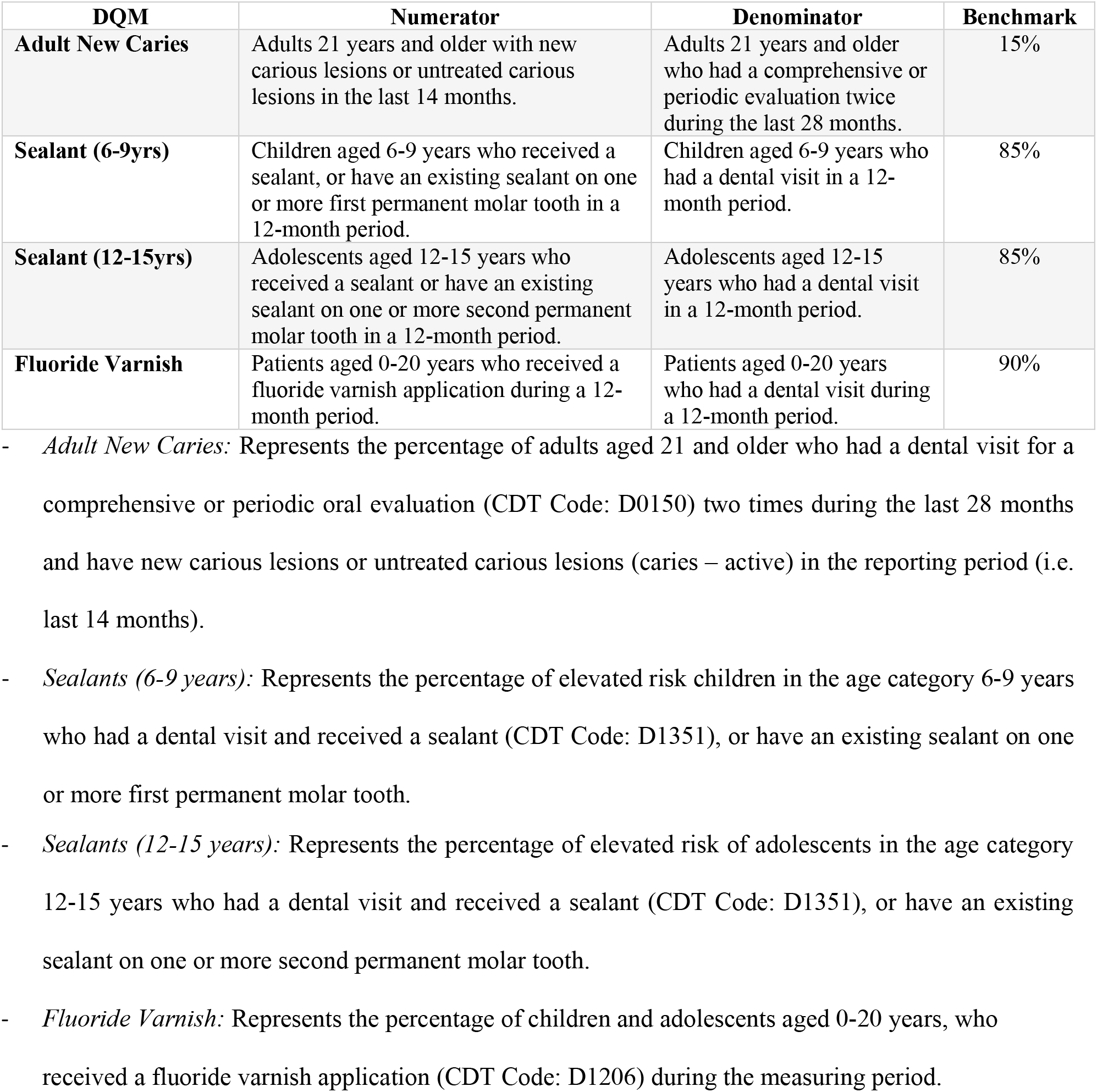
Preventive Dental Quality Measures

### Time Series Modeling

Real-world phenomena captured over time is in the form of a time series. These time series fall under two broad categories. Those whose statistical properties are preserved with time (i.e. stationary) and those whose properties change with time (i.e. non-stationary)^29^. Several models have been proposed in the literature for univariate stationary time series modeling^29,30^. Popular models of stationary time series include moving average models (MA), auto-regressive models (AR) and their combination (ARMA). MA models predict the current value as a linear combination of the past error terms whereas AR models predict the current value as a linear combination of its past and an error term. ARMA essentially model the given time series as a sum of AR and MA models. While ARMA models are useful in predicting stationary time series, statistical properties of DQMs such as those investigated in the present study vary as a function of time rendering them non-stationary. Auto-Regressive Integrated Moving Average (**ARIMA**) are especially well-suited to model non-stationary time series^30^.

### Parameter Estimation and Model Diagnostics

Optimal parameters of ARIMA (p,d,q) with drift were estimated using the Hyndman-Khandakar approach (HK)^31^. KPSS unit root test (α = 0.05)^32^ was used as a part of the HK approach to ensure the stationarity of the integrated time series. Subsequently, maximum likelihood was used to estimate the optimal model parameters and corrected Akaike-Information-Criterion^30^ to penalize model complexity preventing overfitting of the models to the DQM time series. Potential correlation in the residuals obtained as the difference of the original and fitted values were examined using Ljung-Box Portmanteau test (α = 0.05)^33^ as a part of the model diagnostics. The optimal model parameters were subsequently used to generate the 5-month ahead forecasts along with 95% confidence levels^31^. Since the DQMs considered were on different scales, a scale independent measure (**MAPE**: Mean Absolute Percentage Error)^31^ was used to assess the forecasts.

## Results

The time series profiles of the four DQMs sampled at each month from Dec. 2010 to July 2017 is shown in **Fig. 1**. As noted earlier, the first (N = 75) time points were used for estimating the optimal model parameters and the predictive ability of the model along with the 95% confidence level of the predictions was demonstrated on the remaining (N = 5) time points, **Fig. 1**. Adult New Caries exhibited a decreasing trend as a function of time and was modeled using ARIMA (0, 1, 0) with drift. The fitted model along with 5-month ahead prediction and the 95% confidence level are shown in **Fig. 1a**. The corresponding forecast error was 1.8%. Temporal profile of DQM Sealant (6-9 years) exhibited a marginal increase followed by a decreasing trend and was modeled using ARIMA (0, 2, 1). The fitted model along with 5-month ahead forecast and the 95% confidence level are shown in, **Fig. 1b**. The corresponding forecast error was 0.09%. Temporal profile of DQM Sealant (12-15 years) exhibited an increasing trend followed by a marginal decreasing trend and was modeled using ARIMA (0, 2, 1). The fitted model along with 5-month ahead forecast and the 95% confidence level are shown in, **Fig. 1c**. The corresponding forecast error was 0.30%. Temporal profile of DQM Fluoride Varnish exhibited an increasing trend followed by a plateau and was modeled using ARIMA (0, 2, 1). The fitted model along with 5-month ahead forecast and the 95% confidence level are shown in, **Fig. 1d**. The corresponding forecast error was 0.15%. As expected, 95% confidence levels of the predictions increased with time for all the DQMs indicating larger uncertainty in predictions with time.

**Figure 1.**
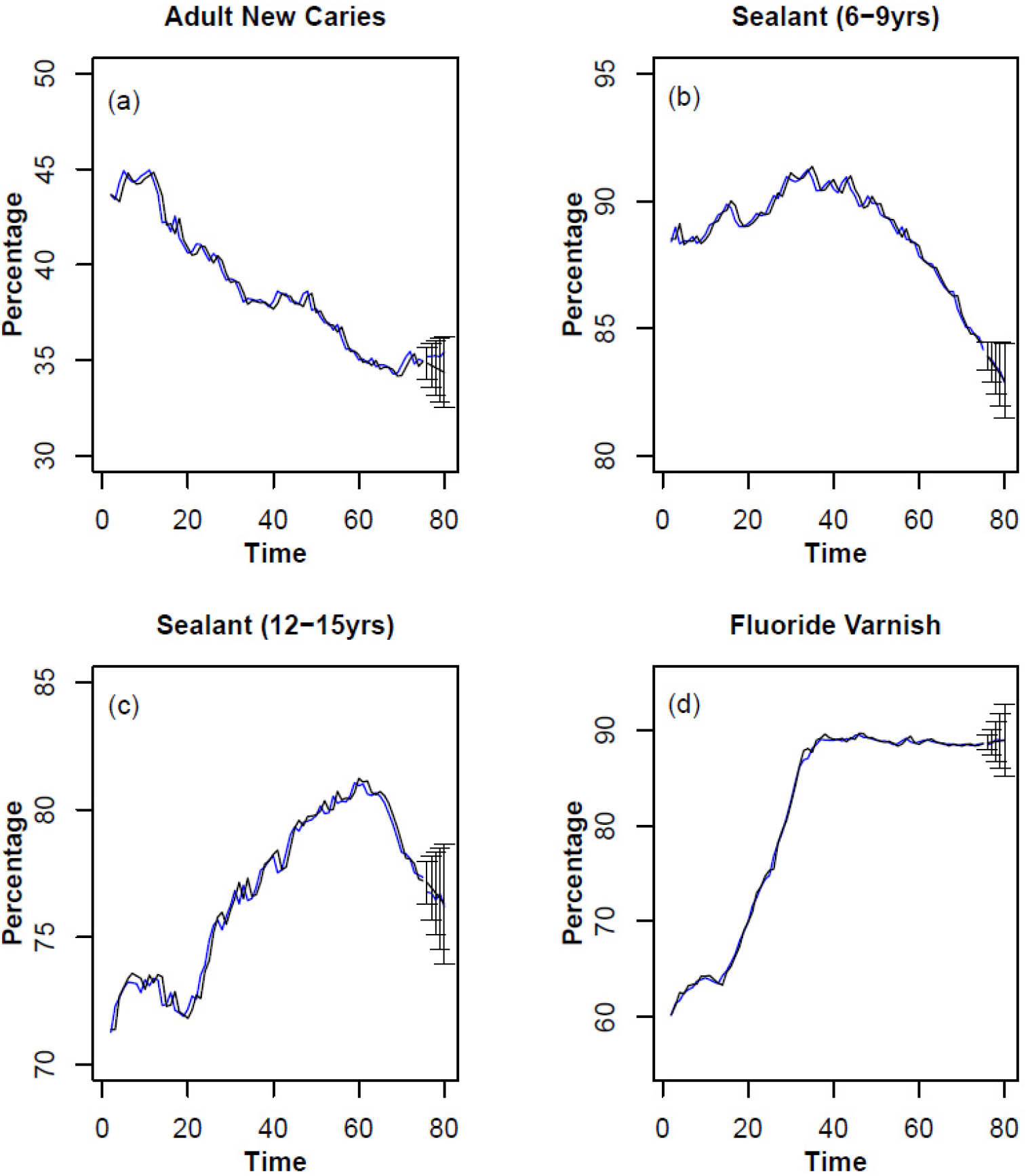
Forecasting DQMs (Adult New Caries, Sealants 6-9 years, Sealants 12-15 years, Fluoride Varnish) is shown in (a-d) respectively. The original monthly time series of the DQMs (blue lines) and the corresponding ARIMA model fit (black lines) along with 5-step ahead forecast and 95% confidence levels (vertical black lines) are also shown in each of the subplots.

## Discussion

Dental quality measures are used routinely to measure system-level performance, self-assessment and sustained performance improvement. While meeting established benchmarks is critical, data-driven forecast as opposed to retrospective review of these preventive measures have the potential to assist in assessing treatment needs ahead of time, strategic decision making, scheduling, allocation of resources and personnel as well as improve oral health literacy initiatives so as improve preventive care utilization and outcomes. More importantly, predictive analytics and forecasting complements traditional descriptive analytics and visualization dashboards^17^. Prediction and forecasting can also assist in prescribing necessary interventions for shifting the DQM trajectories from predicted to desired outcomes ahead of time.

Untreated dental caries is a prevalent chronic disease especially in children with significant economic burden globally^16,19,20^. Therefore, preventive measures for caries is especially critical in rural settings such as those discussed in the present study. These areas are deemed as health professional shortage areas^34^ and face multiple challenges including access to oral health providers, lower insurance rates and oral health literacy challenges. Federally funded safety net programs such as FHCs discussed in the present study have especially been critical in addressing these challenges and improving utilization of preventive services^35^. Dental quality measures are especially helpful in objectively assessing preventive care utilization patterns in rural settings. The present study investigated forecasting preventive DQMs related to caries prevention, including three pediatric measures and one adult measure. Placement of pit and fissure sealants in reducing caries is well documented^22^. Resin-based sealants have especially been shown to reduce caries by (11%-51%) compared to no sealant, when measured at 24 months^36^. The prevalence of sealants in children aged 6-11 years during 2011-2016 in United States was 42%. On an average 3.6 teeth were sealed in children aged 6-11 years with first molar most likely to be sealed^37^. Furthermore the prevalence of sealants in adolescents aged 12-15 years during 2011-2016 in United States was 52%^37^. The first molars were most common teeth sealed in this age group followed by second permanent molars^37^. Forecast modeling of the DQM corresponding to sealants among children (6-9 yrs.) revealed a marginal decrease. While these trends are reassuring^18^, it is important to continue to meet the benchmark (∼85%) in this age group. A more complex trend was observed in the adolescent group (12-15 yrs.), with an initial increasing trend followed by a marginal decreasing trend. The average utilization of sealants in the adolescents (12-15yrs) was historically lower than that of children (6-9 yrs.) in the FHC population, indicating additional efforts in educating the adolescent age groups to get sealants while allocating resources and personnel to attain the benchmark (∼85%). The latter is especially important as dental sealants need to be applied by a dentist, dental hygienist or a qualified dental professional at a dental office or community settings such as school-based sealant programs depending on the state regulations and have been shown to avert caries^38^. Fluoride varnishes have also been shown to minimize incidence of caries in primary and permanent teeth of moderate/high-risk children and adolescents^39^. Water fluoridation has been identified as cost-effective way to reduce caries prevalence^40^. However, community water fluoridation can be a challenge especially in rural areas, as it is not uncommon for communities to use non-fluoridated water sources including private wells^41^. Fluoride varnishes prevent demineralization of enamel, hence helpful in preventing caries. In the FHC patient population, forecast of the DQM corresponding to fluoride varnish showed an increasing trend followed by a plateau indicating proportion of children and adolescents aged 0-20 years seeking fluoride varnish application meeting the benchmark (∼90%). These trends are reassuring and expected to continue based on the forecasts. Prevalence of new adult caries in the FHC’s patient population during the first 75 months of study period showed an encouraging steady decline over the years but the forecast from the models indicated a stabilizing trend that was below the benchmark (∼15%). Thus, it is important to devote sufficient resources for achieving the benchmark. While the present study demonstrated the usefulness of ARIMA time series models in forecasting DQMs at the organizational level, these models may also be used to investigate temporal DQM profiles at center level and those at the level of individual providers with potential assist in determining treatment needs^42^ and sustained performance improvement.

## Data Availability

The data presented in the present study is dental quality measures and can be obtained from the Family Health Center, Marshfield after completing the formalities. The data is also represented as figures in the manuscript.

## Notes

### Competing Interest Statement

The authors have declared no competing interest.

### Funding Statement

The present study is not funded by extramural grants.

### Author Declarations

The study was approved by the MCRI Institutional Review Board.

